# COVID-19 vaccine uptake and associated risk factors among first antenatal care attendees in Zambia, 2021-2022: a repeated cross-sectional study

**DOI:** 10.1101/2024.02.27.24303454

**Authors:** Tannia Tembo, Paul Somwe, Samuel Bosomprah, Kalubi Kalenga, Nyembezi Moyo, Bupe Kabamba, Victoria Seffren, Sombo Fwoloshi, Marie-Reine Rutagwera, Maximillian Musunse, Linos Mwiinga, Julie R. Gutman, Jonas Z. Hines, Izukanji Sikazwe

## Abstract

**Introduction:** Pregnant women are considered a high-risk group for COVID-19, and a priority for vaccination. Routine antenatal (ANC) care provides an opportunity to track trends and factors associated with vaccine uptake. We sought to evaluate COVID-19 vaccine uptake among pregnant women attending ANC in Zambia.

**Methods:** We conducted a repeated cross-sectional study in 39 public health facilities in four districts in Zambia from September 2021 to September 2022. Pregnant women who were aged 15-49 years were enrolled during their first ANC visit. Every month, ∼20 women per facility were interviewed during individual HIV testing and counseling. We estimated vaccine uptake as the proportion of eligible participants who self-reported having received the COVID-19 vaccine.

**Results:** A total of 9,203 pregnant women were screened, of which 9,111 (99%) were eligible and had vaccination status. Of the 9,111 included in the analysis, 1,818 (20%) had received the COVID-19 vaccine during the study period, with a trend of increasing coverage with time (0.5% in September 2020, 27% in September 2022). Conversely, 3,789 (42%) reported not being offered a COVID-19 vaccine. We found that older age, education, employment status, and prior COVID-19 infection were significantly associated with vaccine uptake.

**Conclusion:** COVID-19 vaccine uptake among pregnant women was lower than estimates from the general population (27% across the four districts in September 2022), pointing to missed opportunities to protect this high-risk group. ANC visits were a viable point for conducting COVID-19 surveillance. Incorporating the vaccine as part of the routine ANC package might increase coverage in this group.

**Teaser key message:** Antenatal care clinics could be an easy and sustainable platform for increasing COVID-19 vaccine uptake among pregnant women.

**Key Findings:**

- Despite evidence that COVID-19 vaccines are safe in pregnancy, vaccine uptake was low among pregnant women in Zambia through the end of 2022.
- Various demographic characteristics were associated with the uptake of COVID-19 vaccines.

**Key Implications:**

- It is acceptable and feasible to assess COVID-19 vaccination uptake and hesitancy among pregnant women attending ANC. ANC may provide an easy, sustainable platform for routinely monitoring COVID-19 and other disease outbreaks.
- Key vaccination messages and mass vaccination strategies can be designed and adopted for pregnant women to reduce vaccine hesitancy and consequently, increase vaccine uptake.
- COVID-19 vaccination could be incorporated into routine antenatal care to help increase vaccination coverage among pregnant women seeking antenatal care services.

## INTRODUCTION

The coronavirus disease 19 (COVID-19) pandemic has presented unprecedented challenges globally, affecting all aspects of life, including the health of vulnerable populations. As of October 2023, there were approximately 771,151,224 confirmed cases of COVID-19 and 6,960,783 deaths, globally. ^1^ Due to immunologic and physiologic changes, pregnant women are considered a high-risk group for COVID-19. ^2–4^ Those with SARS-CoV-2 infection are more susceptible to severe disease and mortality as compared to non-pregnant women. ^5,6^ Although COVID-19 vaccines are recommended for pregnant women to reduce the risk of severe COVID-19 disease and prevent adverse obstetric outcomes, vaccine hesitancy – a delay or refusal of safe vaccines– is reported to be higher among pregnant women than in the general population. ^7,8^ While survey results from Iran show that more than two-fifths of participants accepted receiving any vaccine, ^9^ fewer than one-third of pregnant women were interested in receiving the COVID-19 vaccine in Cameroon. ^10^

Understanding the factors influencing COVID-19 vaccine uptake among women attending antenatal care (ANC) is crucial for mitigating the impact of the virus on maternal and child health. Previous studies have shown variations in COVID-19 vaccine uptake among different populations, highlighting the influence of various factors such as age, education level, and socioeconomic status. ^11–13^ However, there is a limited understanding of these factors, particularly among pregnant women, who have distinct healthcare needs and considerations. Vaccination not only protects pregnant women from severe illness but also offers potential benefits to their unborn children through passive immunity. ^14,15^ By identifying risk factors and barriers to vaccine uptake, we can develop tailored interventions that address specific concerns and enhance vaccine acceptance, ultimately safeguarding the health and well-being of both expectant mothers and their babies. In Zambia, COVID-19 vaccines were first made available in April 2021. The voluntary vaccination exercise initially targeted people above the age of 18 years and prioritized frontline health workers, people involved in the maintenance of core societal function, people with underlying conditions, and those in congregate settings. Despite initial uncertainties about the safety of COVID-19 vaccines in pregnancy, the Ministry of Health (MoH) updated guidelines in July 2021 to include vaccination for pregnant women. ^16^ Vaccine uptake among pregnant women in Zambia remains unknown as available evidence is based on results of vaccine acceptance among the general population. ^17,18^

This study aims to estimate COVID-19 vaccine uptake and associated risk factors among pregnant women attending their first antenatal care (ANC) visits at public health facilities in Chadiza, Chipata, Chongwe, and Lusaka districts of Zambia during the height of the third (Delta variant) and fourth (Omicron variant) waves. We also sought to assess the feasibility of using routine ANC to track trends and factors associated with the uptake of vaccines. This study contributes to a broader knowledge base on COVID-19 vaccination by focusing on specific population subsets, which can provide useful guidance on the development of targeted communication campaigns, healthcare policies, and interventions aimed at improving vaccine uptake and reducing disparities within this vulnerable group.

## MATERIALS AND METHODS

### Study Design

We conducted repeated cross-sectional surveys on SARS-CoV-2 seroprevalence and vaccination coverage in pregnant women attending their first ANC visit in the maternal, newborn, and child health (MNCH) clinics at 39 rural and urban public health facilities in four districts (Chadiza, Chipata, Chongwe, and Lusaka). Participant recruitment was initiated on 4 September 2021 and was concluded on 30 September 2022 for study sites in Chipata, Chongwe and Lusaka districts. Initiation of participant enrolment was delayed at study sites in Chadiza and only started on 23 September 2021 and ended on 31 July 2022. In Zambia, the health system is classified into three categories: 1) community-level health posts and health centers; 2) Level 1-provincial and district hospitals and 3) Level 2-central-level specialized hospitals. ^23^ Study sites (all public health facilities) were stratified according to location and selected using probability proportional to facility size based on each health facility’s monthly mean number of pregnant women attending their first ANC visit based on historic monthly ANC attendance during the 2020 fiscal year. Study activities were incorporated into routine ANC services and leveraged infrastructure and human resources in the MNCH departments at public health facilities. In Chadiza district, study activities were incorporated into an ANC-based malaria surveillance pilot study. Health facility staff (CHWs, midwives, and phlebotomists and research assistants were trained in the study protocol, laboratory procedures, and human subject protection. Additional methodological details of the study are described elsewhere. ^19^ This analysis looks at interview data on COVID-19 vaccination from the broader study.

### Recruitment and Enrollment of Study Participants

A CHW provided information on the ANC COVID-19 surveillance study during sensitization meetings in the community and group counseling sessions at the health facility. Once every week, on a day that each health facility provided routine ANC services for first-time attendees, CHWs conveniently identified pregnant women and screened them for eligibility to participate in the study. Pregnant women who were aged 15-49 years, confirmed to be pregnant, registered as a first ANC attendee, and provided consent, were enrolled in the study. Potential participants were screened for active SARS-CoV-2 infection using a paper-based checklist of symptoms. Those who showed one or more symptoms were excluded and referred to a SARS-CoV-2 testing center to be managed according to national guidelines if they had a positive test result. At each study site, trained midwives shared an information sheet with eligible participants and obtained consent from up to 20 pregnant women per month, cumulatively enrolling 200 pregnant women per district.

### Data Collection and Management

At the time of providing routine ANC tasks such as performing physical examinations, administering antimalaria prophylaxis, prescribing prenatal vitamins and drawing blood samples for routine HIV and syphilis testing, a midwife administered an electronic questionnaire to consented participants. The study questionnaire was designed in Open Data Kit (ODK) – a free, open-source suite of software tools that facilitates the collection, management and use of data using portable mobile devices with Internet connectivity. ^24^ To ensure confidentiality and privacy, a unique identification number was assigned to individual participants during data collection. The questionnaire was uploaded onto a tablet which was handled by trained midwives and research assistants.

To estimate COVID-19 vaccine uptake and associated risk factors among pregnant women attending their first antenatal care, the study collected participants’ vaccination status, socio-demographic characteristics, self-reported past SARS-CoV-2 infection, and exposure. SARS-CoV-2 preventive measures and behavior such as the proper wearing of masks were observed and captured in the questionnaire. In addition, the study determined the acceptability and feasibility of monitoring COVID-19 vaccination uptake and hesitancy among pregnant women attending ANC. Acceptability was defined as a multi-faceted construct that reflects the extent to which the healthcare providers and pregnant women enrolled in the study considered the SARS-CoV-2 surveillance to be appropriate and was measured by its perceived effectiveness – the extent to which the SARS-CoV-2 surveillance was perceived as likely to achieve its purpose. ^20^ Feasibility was defined as the study’s appropriateness for further testing to determine relevance and sustainability ^21^ and was measured by recruitment rates, length of time from initiation to completion of the targeted number of participants, and number of eligible participants required to recruit sample size.

Initially, participants were asked only if they had been vaccinated or not. The questionnaire was updated in May 2021 to gather information about whether the participants had been offered a vaccine in addition to whether they had accepted it, and for those who were unvaccinated, additional information was collected about the reasons for not being vaccinated. Overall, 3,655 participants were recruited before the updated questionnaire was implemented, meaning they only had a binary vaccination status collected. COVID-19 vaccination status was self-reported, although study staff attempted to verify information from vaccination cards if participants had them available on the day of their ANC visit.

Data were reviewed on a scheduled basis and missing variables were communicated to health facility staff and research assistants to attempt follow-up and completion. All completed questionnaires were uploaded onto a central server individually or batched. Other data were documented in study-specific logs and registers. Additionally, COVID-19 vaccine coverage data in the general population in the four implementation districts were obtained from the Zambia MoH so that a comparison could be made to the estimates from this study. General population vaccination data were extracted from the national District Health Information System (DHIS) 2-COVAX tracker – a global digital health data toolkit that facilitated the capture of individual-level COVID-19 data of investigated cases; including vaccination status. ^22^

### Statistical Analysis

A sample size of 200 women per district per month was selected based on estimating an acceptable margin of error for varying SARS-CoV-2 seroprevalences that were expected to vary by location and enrollment month (this sample size would produce a 95% confidence interval [CI] half-width of 2.4% at the lowest expected seroprevalence of 3%). The sample size was adjusted to account for a design effect of 1.5. Background characteristics were summarized using frequency and proportions for categorical variables, and median and interquartile range for continuous variables. We estimated the prevalence of COVID-19 uptake as the proportion of pregnant women who received (i.e., reported or verified by vaccination card) COVID-19 vaccine.

The corresponding 95% CIs were adjusted for the clustering of participants within a health facility. We used a mixed-effect logistic regression model to identify factors independently associated with uptake. To allow for variation in the secular trend across clusters (i.e., health facilities), we extended the random-effects components to allow a random interaction between time and health facilities (i.e., we generated a new variable by combining two dimensions of health facilities and time). P-values less than 0.05 were considered statistically significant. All statistical analyses were performed using Stata 18 MP (StataCorp, College Station, TX, USA).

### Ethics Considerations

This study was reviewed and approved by the National Health Research Authority (NHRA), a statutory body under the Zambian MoH, and following the US Centers for Disease Control and Prevention (CDC) human research protection procedures as the CDC investigators were not engaged (i.e., did not interact with human subjects or have access to identifiable data or specimens for research purposes). It was also reviewed and approved, particularly for the Chadiza sites, by the University of Zambia Biomedical Research Ethics Committee (UNZABREC) and PATH’s Research Ethics Committee (PATH REC). Information sheets for participants were developed and translated into two local languages. Individual written consent and assent were obtained from adults and minors aged below 18 years, respectively, in a language of their choice and a copy of an information sheet was offered to the participant after consenting. Assent was obtained from minors only after consent was given by their parent(s) or guardian(s). Illiterate adults were required to provide consent in the presence of an impartial witness.

## RESULTS

### Study Enrollment Cascade

During the study period, a total of 9,203 pregnant women were approached to participate in the study. Of these, 19 (0.2%) were ineligible due to their age and were excluded (Figure 1). A further 73 did not have vaccination status, giving a total of 9,111 included in the analysis. Overall, 1,818 (20.0%) pregnant women were vaccinated against COVID-19 by the time of their first ANC visit and, of 7,293 unvaccinated women, 5,456 had additional vaccination information collected (i.e., were administered the updated questionnaire). At least 1,667 (30.6%) had been offered a vaccine but refused (Figure 1) and provided reasons for refusal (Figure 3).

**Figure 1:**
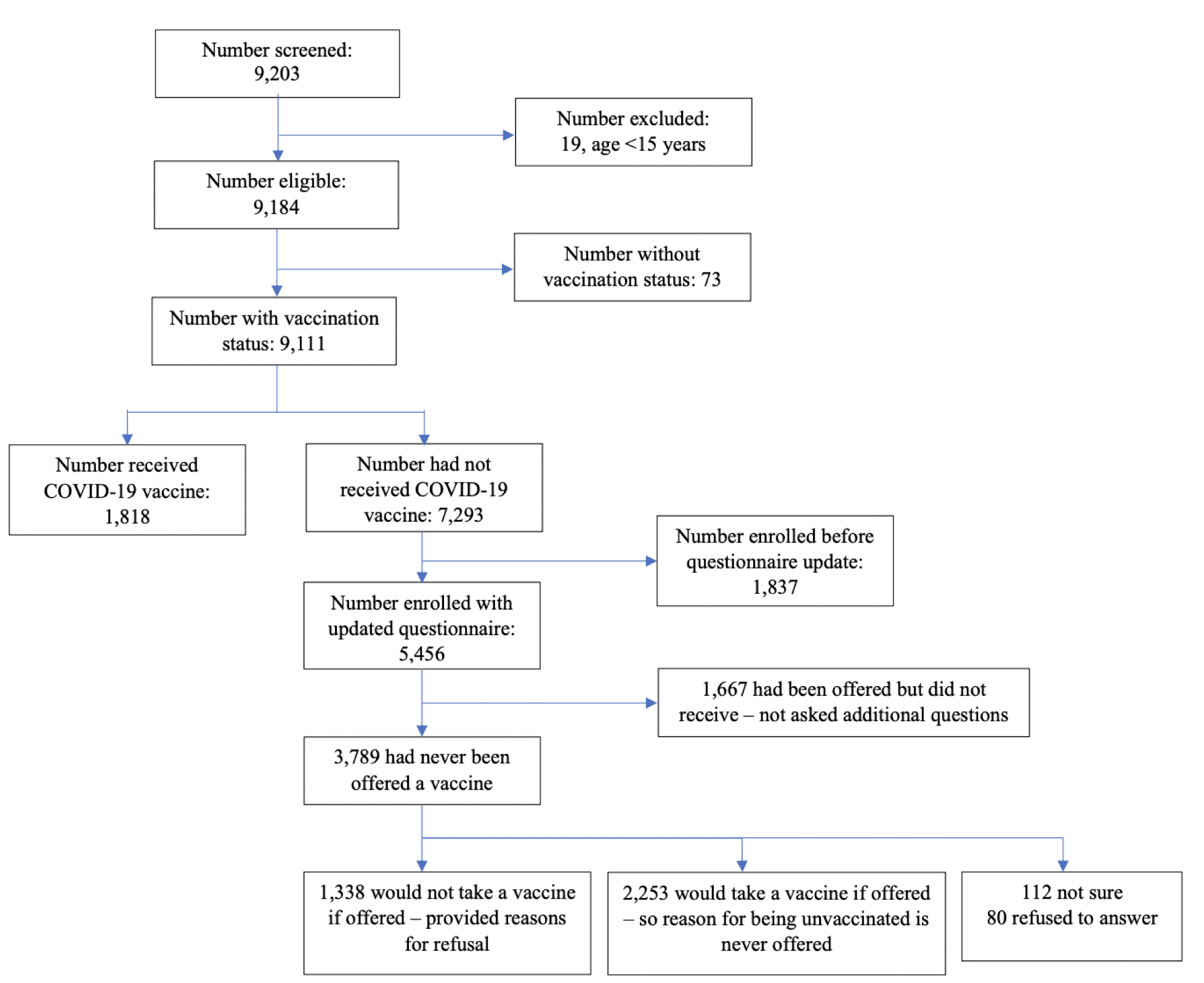
Flow chart of participant recruitment.

### Background Characteristics and COVID-19 Uptake

The median age of participants was 24 years (interquartile range [IQR]: 20-30) and over half (4,944 [54.3%]) of participants were aged between 20-29 years (Table 1). Half of the participants (4,557) had attained some or completed primary school education while half were employed either formally or informally. Almost all, (8,917 [95%]) participants had never tested for COVID-19 before their ANC visit (Table 1). An estimated 4% of pregnant women reported ever being in contact with a COVID-19-infected individual, either within and/or outside their household. Proper use of masks was observed in only 3,492 (38.3%) of the participants. Seven hundred ninety-one (8.7%) of the pregnant women either tested positive for HIV or had a known positive HIV status on the day of their visit. Of the 9,111 participants with a vaccination status, 1,818 (20%) reported having received a COVID-19 vaccine while 3,789 (41.6%) of women said they had never been offered a vaccine (Figure 1).

**Table 1.**
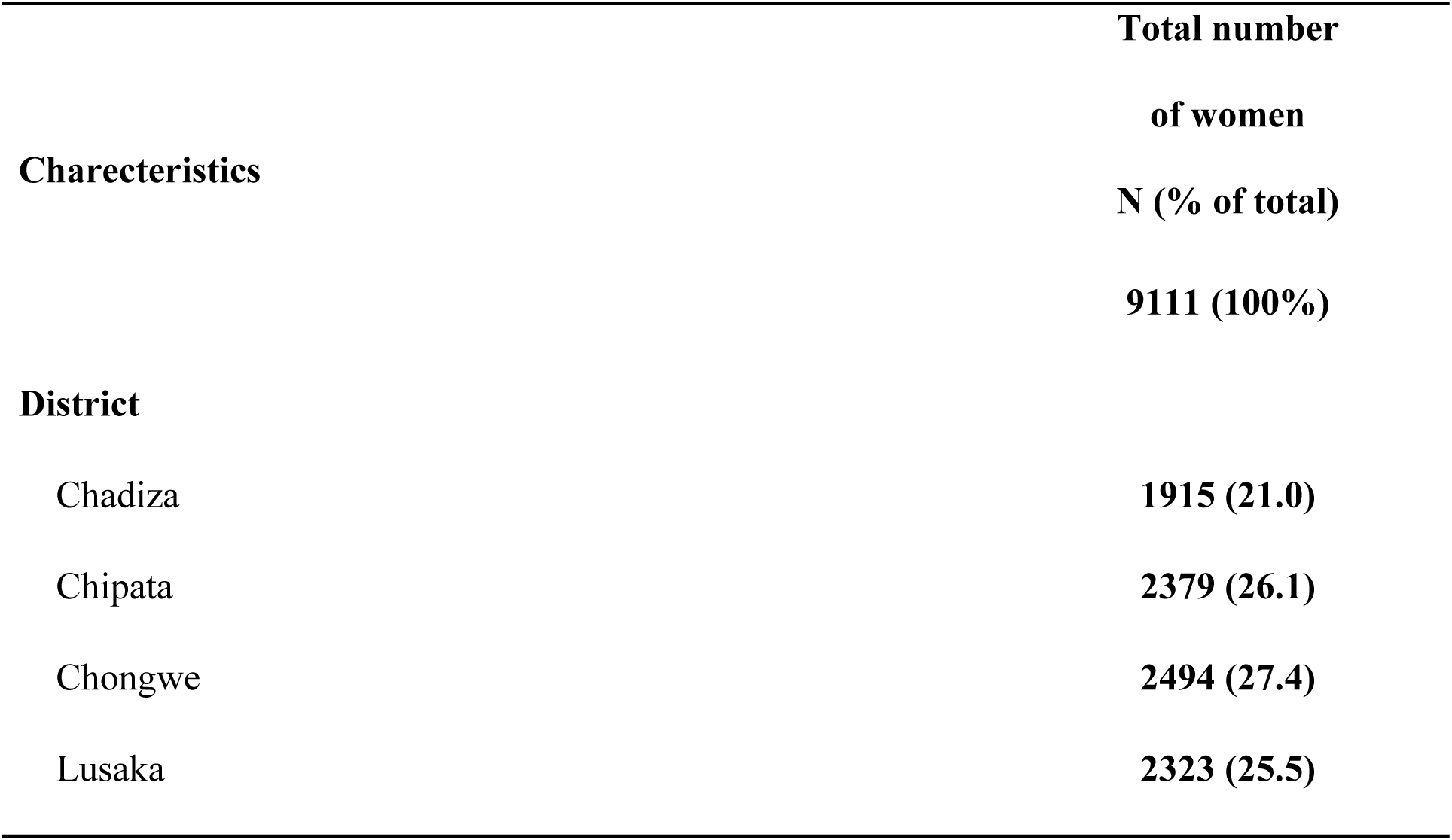
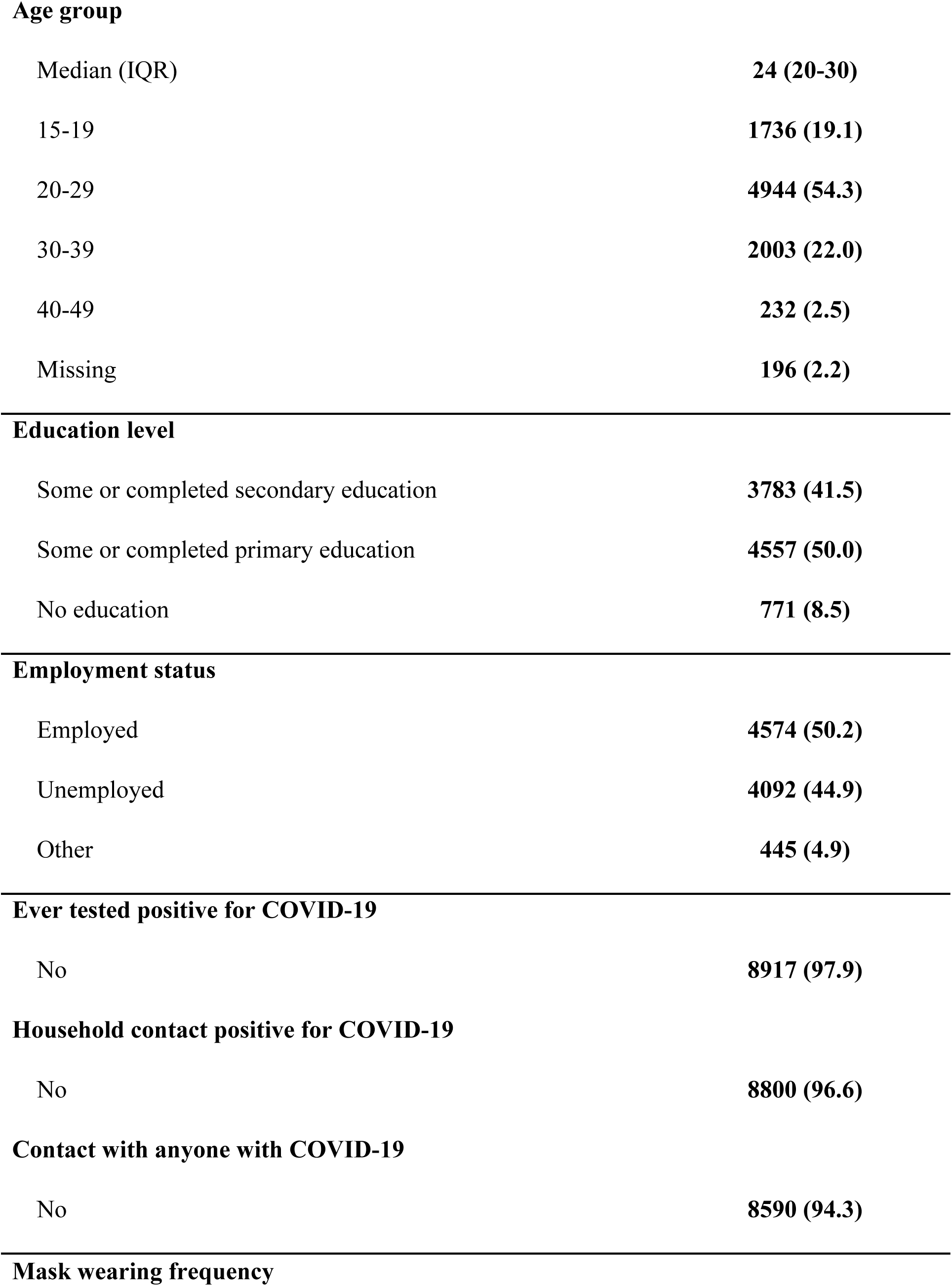
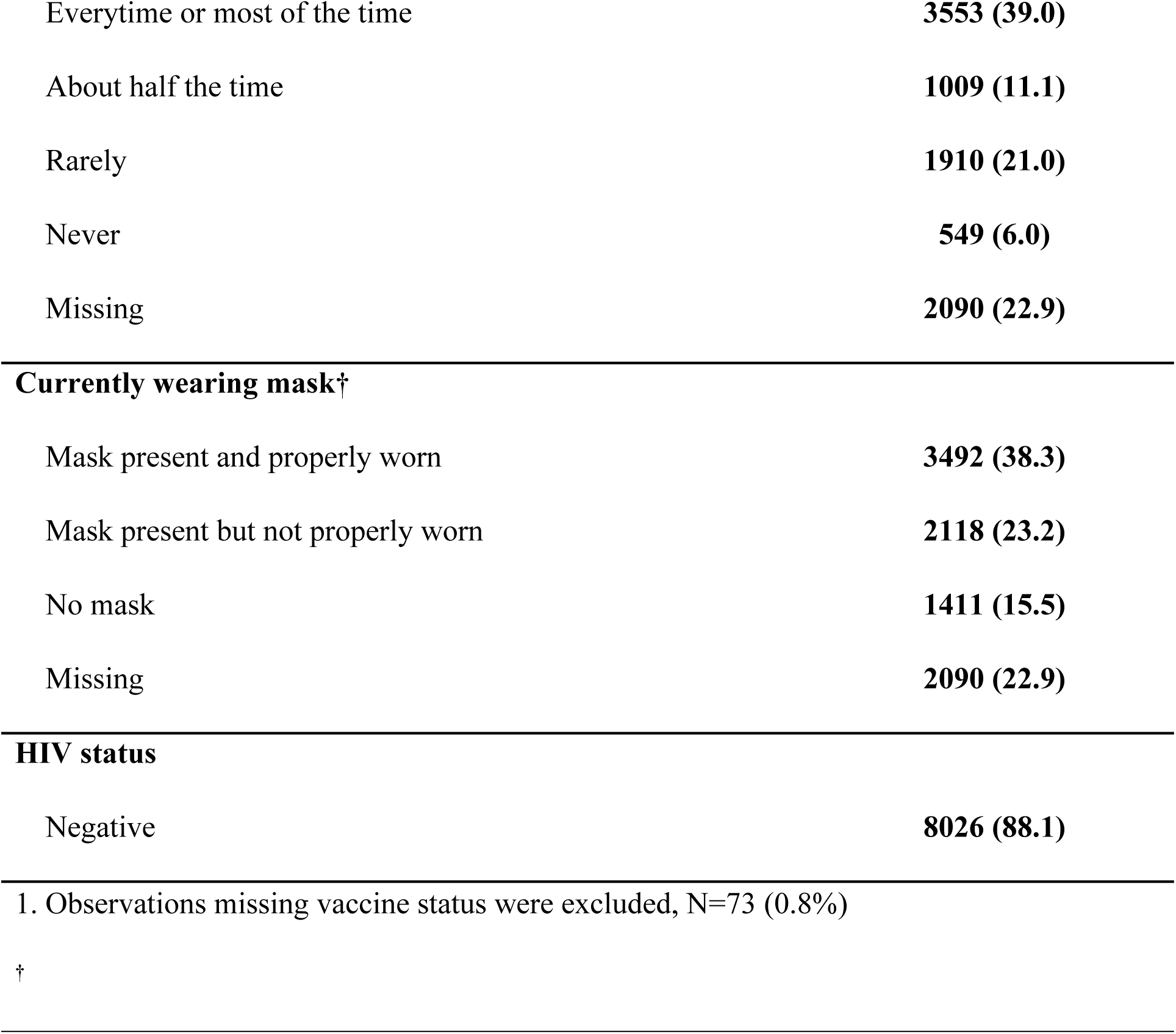
COVID-19 vaccine uptake among pregnant women attending first ANC visit by socio-demographic characteristics, N=9111.

| **Table 1. Background characteristics and COVID-19 vaccine uptake among pregnant women attending first ANC visit** |  |
| --- | --- |
| **Charecteristics** | **Total number**<br>**of women**<br>**N (% of total)** |
|  | **9111 (100%)** |
| **District** |  |
| Chadiza | **1915 (21.0)** |
| Chipata | **2379 (26.1)** |
| Chongwe | **2494 (27.4)** |
| Lusaka | **2323 (25.5)** |
| **Age group** |  |
| Median (IQR) | 24 (20-30) |
| 15-19 | 1736 (19.1) |
| 20-29 | 4944 (54.3) |
| 30-39 | 2003 (22.0) |
| 40-49 | 232 (2.5) |
| Missing | 196 (2.2) |
| **Education level** |  |
| Some or completed secondary education | 3783 (41.5) |
| Some or completed primary education | 4557 (50.0) |
| No education | 771 (8.5) |
| **Employment status** |  |
| Employed | 4574 (50.2) |
| Unemployed | 4092 (44.9) |
| Other | 445 (4.9) |
| **Ever tested positive for COVID-19** |  |
| No | 8917 (97.9) |
| **Household contact positive for COVID-19** |  |
| No | 8800 (96.6) |
| **Contact with anyone with COVID-19** |  |
| No | 8590 (94.3) |
| **Mask wearing frequency** |  |
| Everytime or most of the time | **3553 (39.0)** |
| About half the time | **1009 (11.1)** |
| Rarely | **1910 (21.0)** |
| Never | **549 (6.0)** |
| Missing | **2090 (22.9)** |
| **Currently wearing mask<sup>†</sup>** |  |
| Mask present and properly worn | **3492 (38.3)** |
| Mask present but not properly worn | **2118 (23.2)** |
| No mask | **1411 (15.5)** |
| Missing | **2090 (22.9)** |
| **HIV status** |  |
| Negative | **8026 (88.1)** |

1. Observations missing vaccine status were excluded, N=73 (0.8%)
<sup>†</sup>

### Factors independently associated with vaccine uptake

In a multivariable mixed-effects logistic regression model, we found that age group, education, employment status, ever testing positive for COVID-19, and HIV infection were all significantly associated with COVID-19 vaccine uptake (Table 2). Increasing age was associated with increased COVID-19 vaccination uptake. We observed a lower proportion of COVID-19 vaccination among pregnant women who had no education ((aPR): 0.79, 95% CI: 0.62-1.00; P = 0.007) or had attained some or completed primary education (aPR): 0.86 95%CI: 0.75-0.97; P = 0.001) as compared to those who had some or completed secondary education. Unemployed pregnant women had lower vaccination prevalence ((aPR): 0.69, 95% CI: 0.58-0.82; P = <0.001) as compared to pregnant women who were employed. Additionally, pregnant women who had ever tested positive for COVID-19 had 1.70 times (95% CI: 1.31-2.21; P = <0.001) greater likelihood of receiving a vaccine compared to those who had not tested positive for COVID-19 in the past. Similarly, pregnant living with HIV were 1.18 times (95% CI:1.02-1.37; P = 0.026) more likely to be vaccinated compared to those who were not infected with HIV.

**Table 2.**
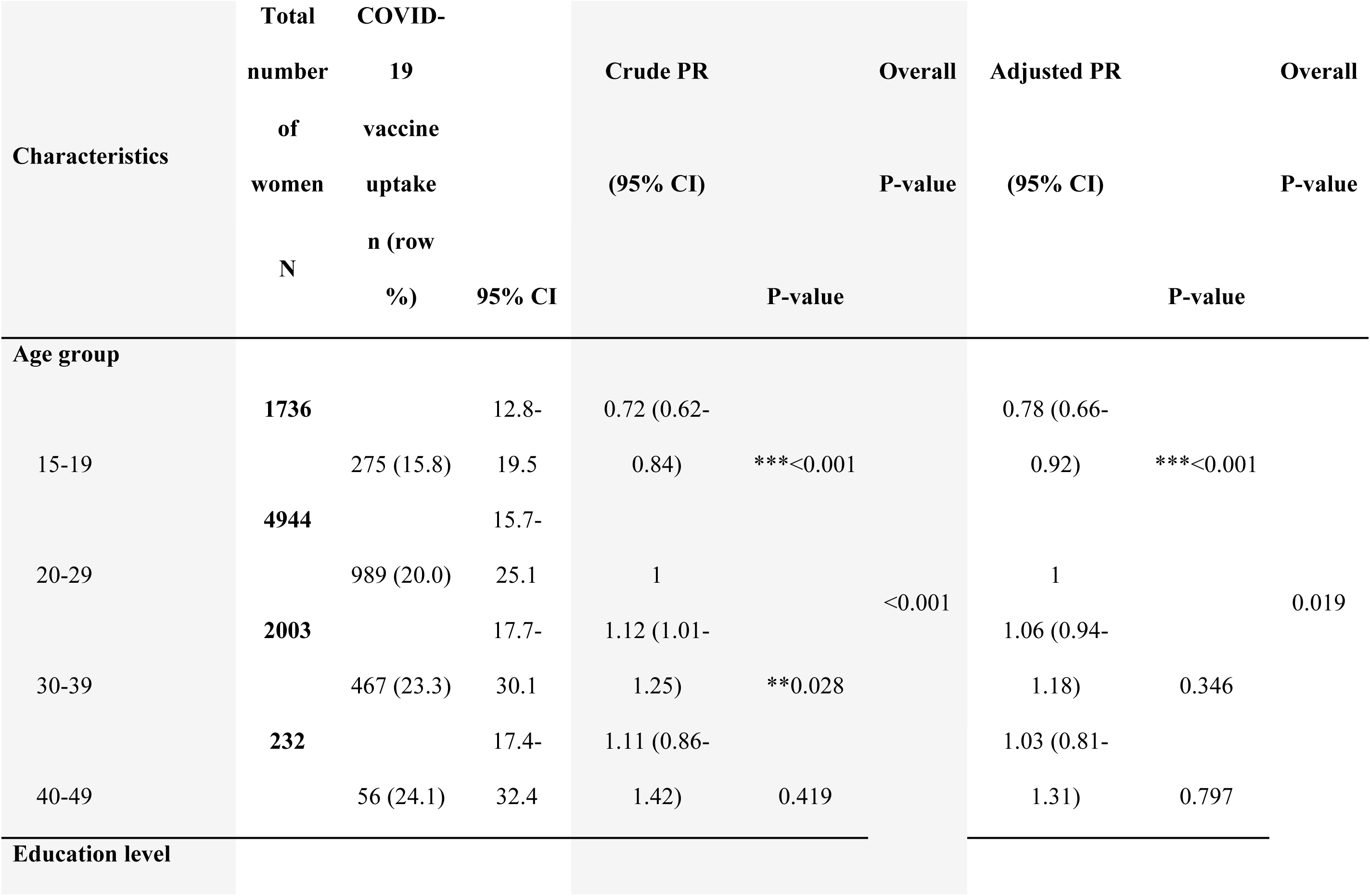
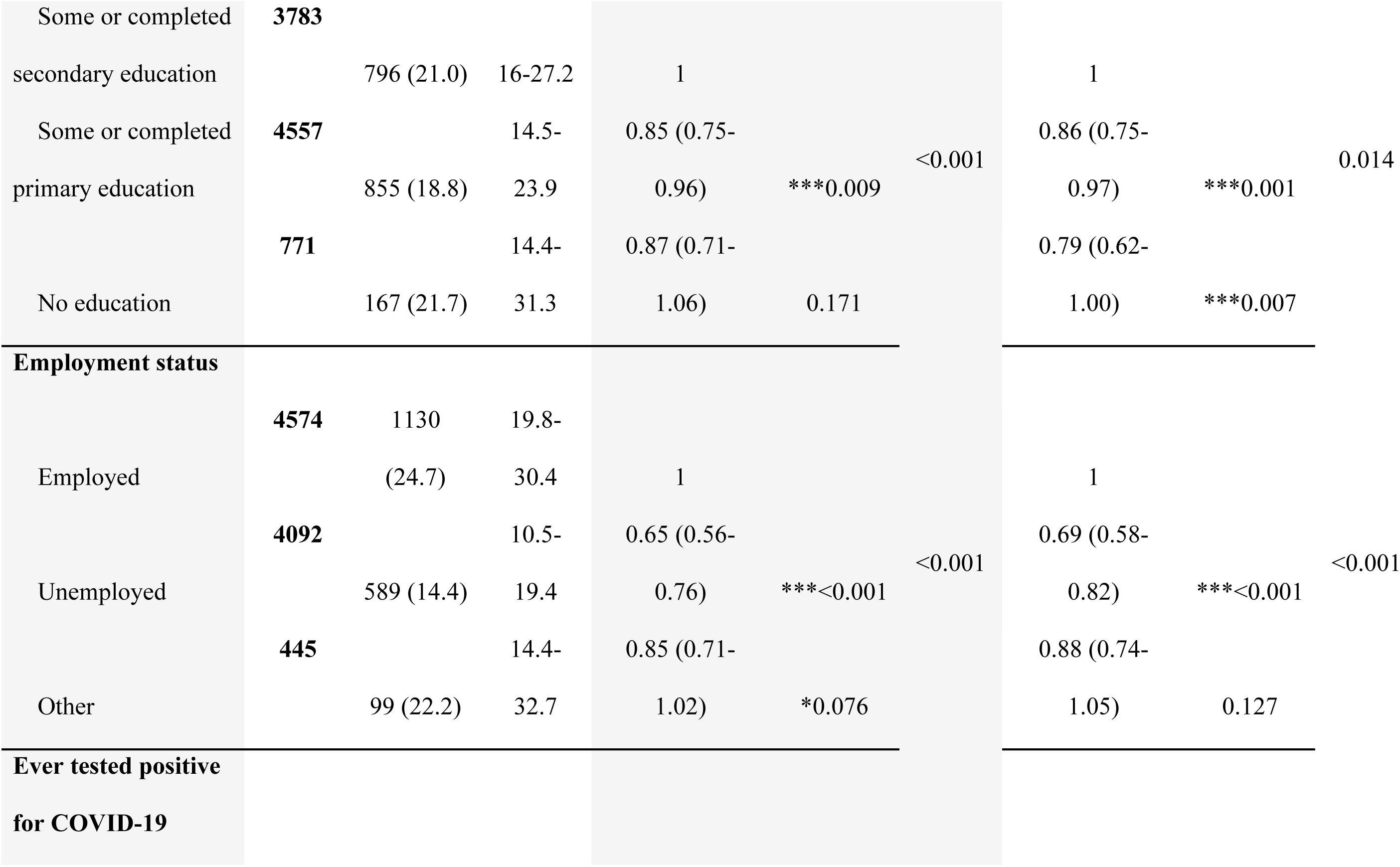
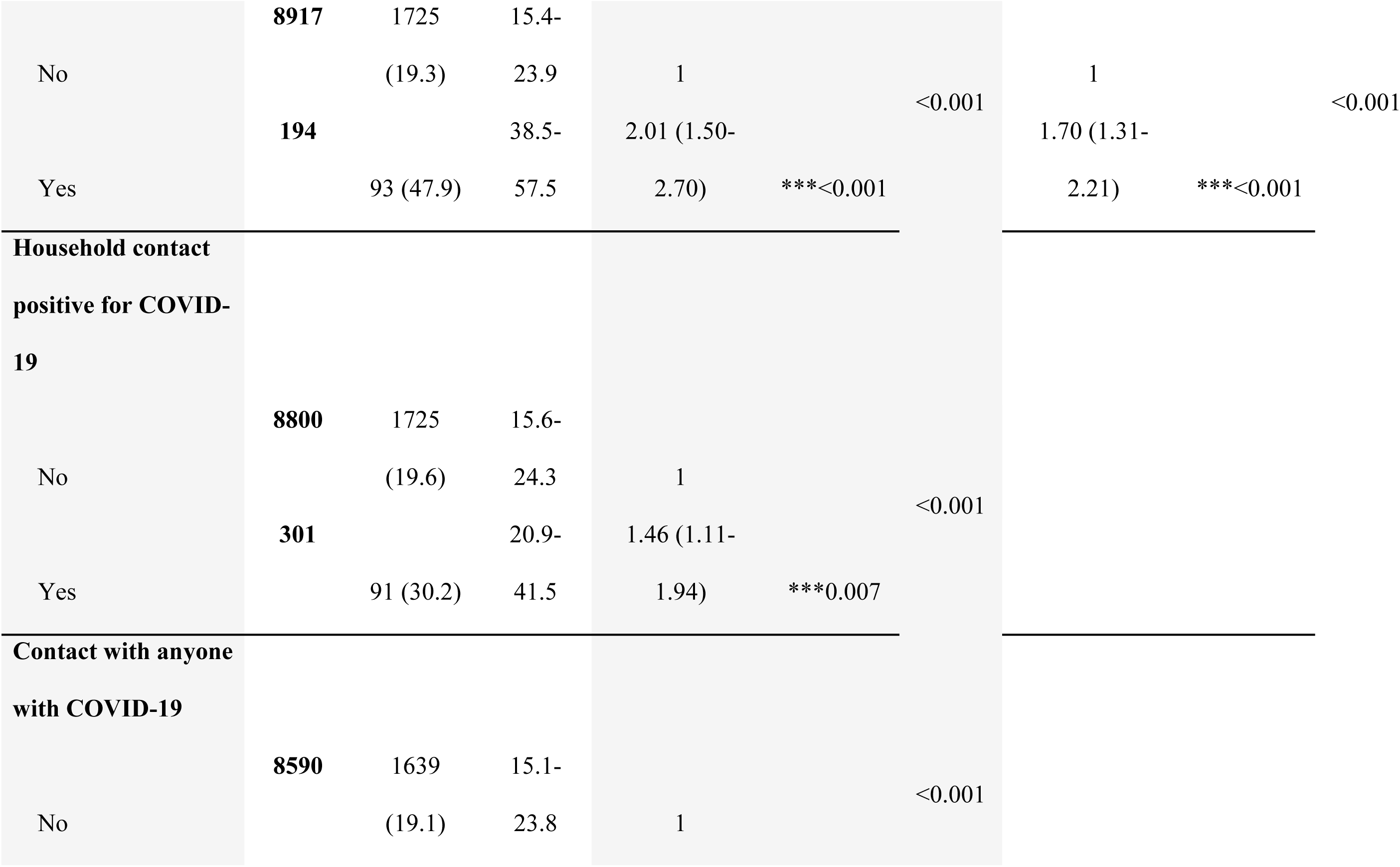
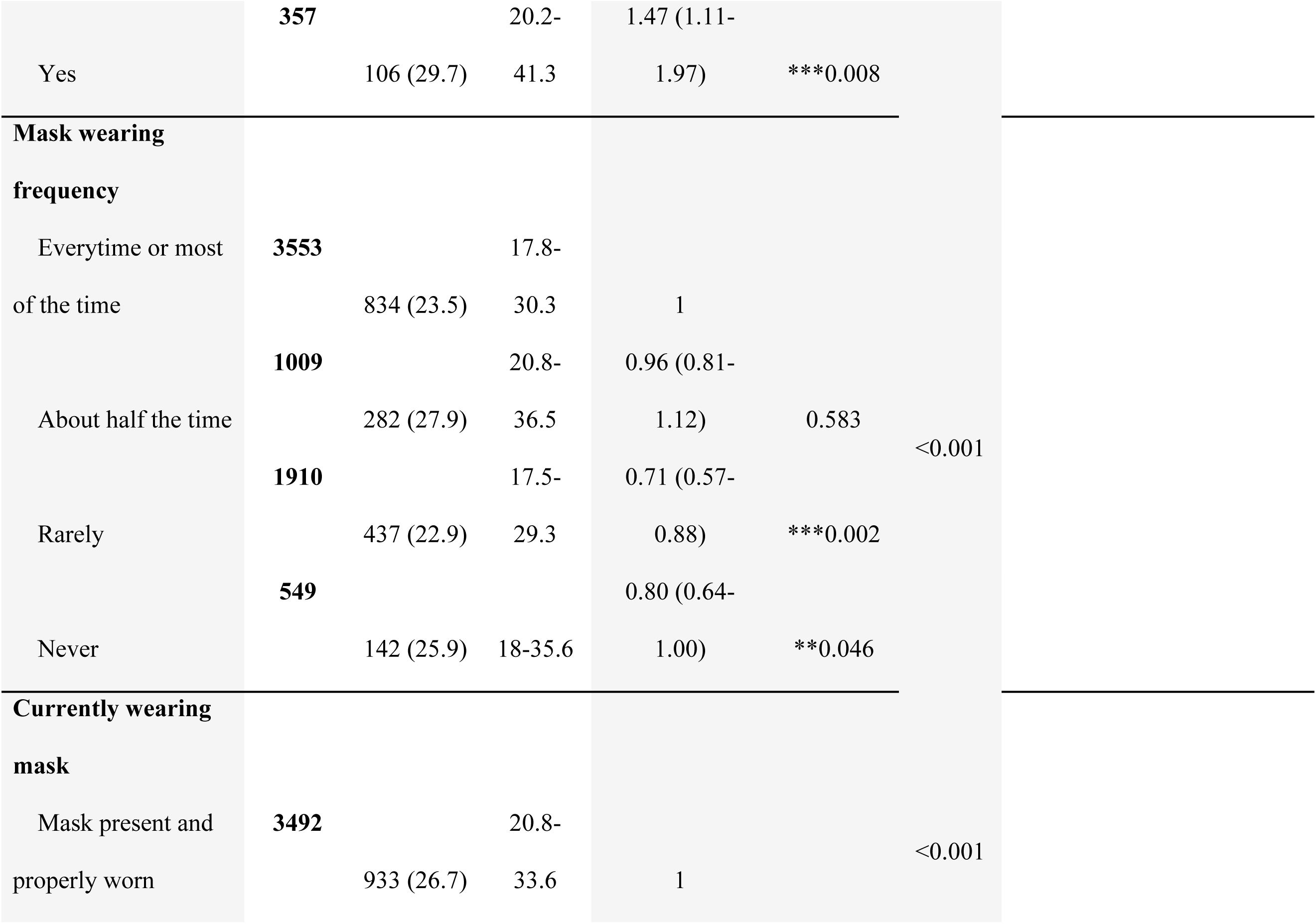
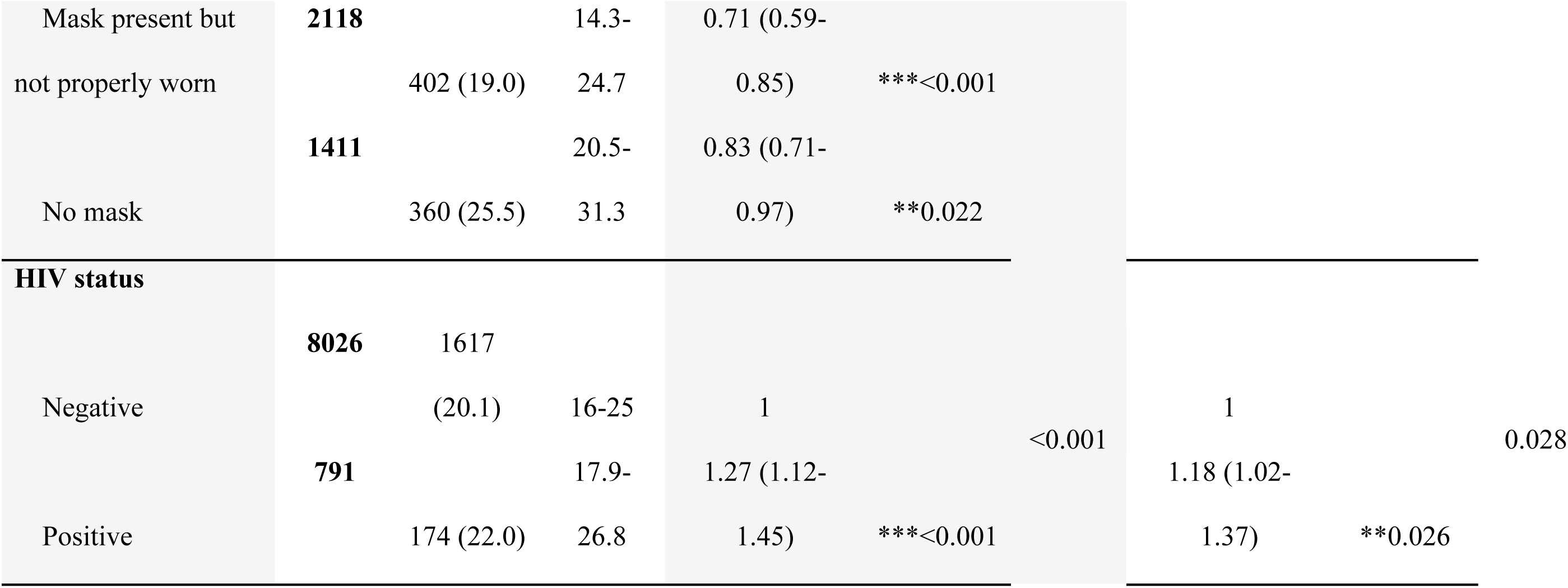
Factors associated with vaccine coverage among pregnant women attending first ANC visit, n=8,750.

### Vaccine uptake over time

Overall, COVID-19 vaccination uptake among pregnant women was at 0.5% at the beginning of the study in September 2021 and had increased to 27% by September 2022 (Figure 2). When compared by district, uptake varied between 2.8% and 17.4% in September 2021 and 21.8% to 29% in September 2022. Throughout implementation, a steady increase in vaccine uptake was observed among participants in Chadiza (Figure 2). COVID-19 vaccine coverage among women at first ANC visits was lower than coverage in the general population in these districts (Figure 2).

**Figure 2:**
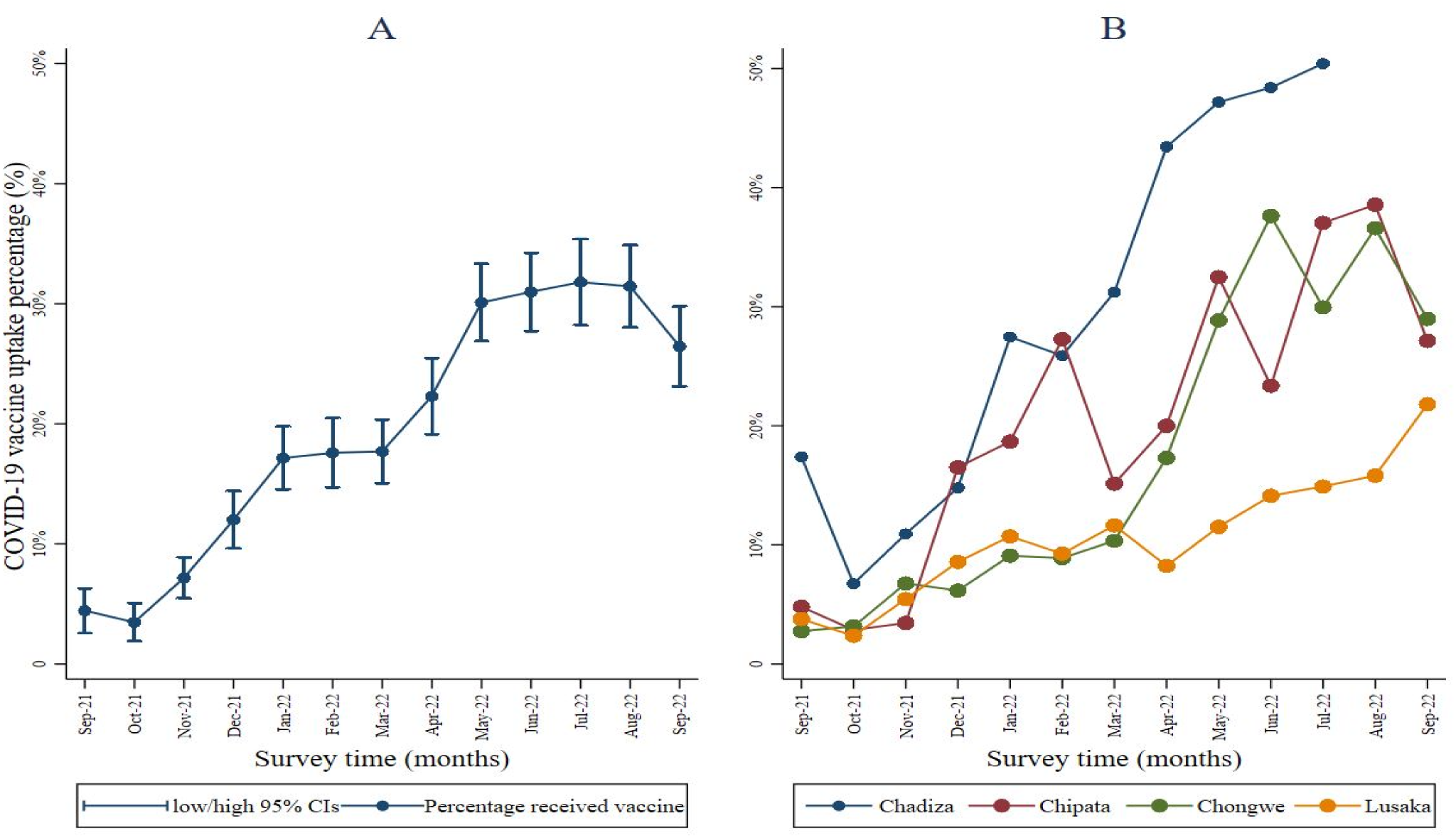
Vaccine uptake among pregnant women by facility (September 2021 to September 2022, N=9,111)

**Figure 2.**
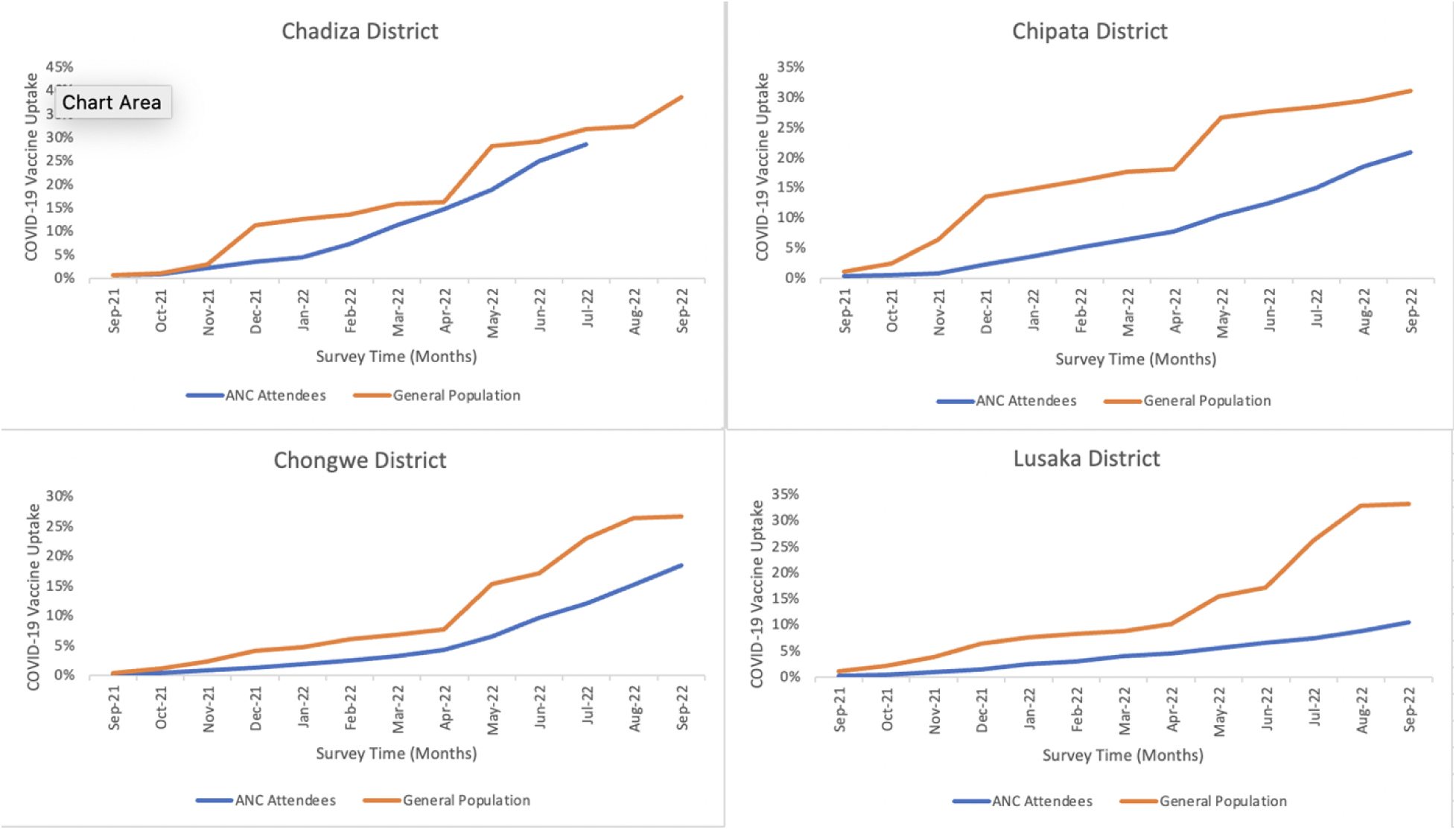
COVID-19 vaccine coverage estimates in pregnant women attending first ANC visit compared with estimates in the general population.

**Figure 3:**
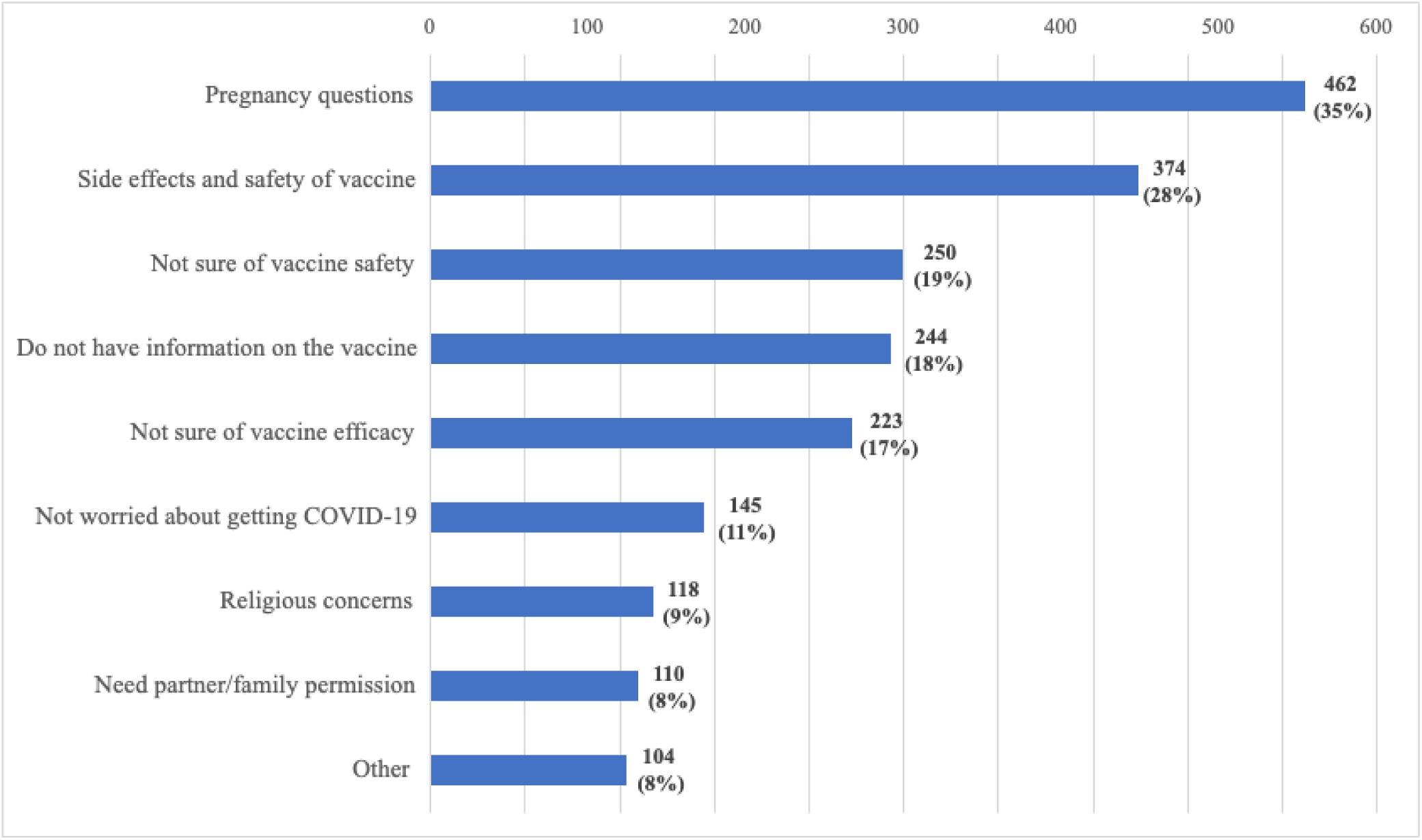
reasons for not taking vaccine if offered, n=1,338.

### Reasons for not taking the COVID-19 vaccine if offered

Of the 3,789 pregnant women who reported never having been offered a vaccine, 1,338 (35.3%) said that they would not take a COVID-19 vaccine even if they were offered and provided reasons for refusal (Figure 3). Pregnancy safety concerns and vaccine effects were the most common reasons for refusing to take the vaccine (462 [35%] and 374 [28%], respectively). An additional 250 (19%) would not take it because of uncertainties surrounding its safety, and 244 (18%) of the participants would not take a vaccine because they did not have sufficient information on the vaccine. The least cited reasons for refusal to take a COVID-19 vaccine were religious concerns and the need to seek permission from a partner or family (9% and 8%, respectively).

## DISCUSSION

Most pregnant women in the four districts of Zambia were not vaccinated against COVID-19, despite being at high risk for severe outcomes. Vaccination coverage increased over the study period and was higher in all four districts compared to contemporaneous coverage estimates of the general population. Despite evidence that COVID-19 vaccines are safe in pregnancy, only ten of the 54 African countries were recommending COVID-19 vaccines for pregnant women as of February 2022. ^23^ In Zambia, COVID-19 vaccines were first made available for the general population in April 2021 ^24^ but, vaccine coverage remains well below the World Health Organization’s (WHO) target to vaccinate 70% of the world’s population by mid-2022. ^25^ Given initial uncertainty about vaccine safety in pregnancy, the MoH delayed recommending COVID-19 vaccination for pregnant women until July 2021, which might have contributed to vaccine hestitancy in this group. Our findings correlate with a study conducted in Ethiopia in which only 14.4% of the participants had received at least one dose of COVID-19 vaccines by March 2022. ^26^ Similarly, vaccine uptake in Cameroon was low (31%) but, uptake did not differ between the pregnant and general population. ^27^

Low vaccination rates in low-and-middle-income (LMICs) African countries are partly due to the inequitable distribution of vaccines caused by constrained financial resources to support competing priorities in underdeveloped health systems and bottlenecks in the supply chain which may lead to delayed vaccine rollout. ^28,29^ Studies have also reported that pregnant women did not receive COVID-19 vaccines due to their limited availability in Africa, perhaps an indication of the inability of pharmaceutical companies to manufacture vaccines in Africa, and the compromised quality of the vaccines manufactured by international pharmaceutical companies for the African market. ^27,30^ COVID-19 vaccination uptake is also hampered by the low-risk perception of the pandemic, concern about adverse vaccine effects, low vaccination awareness, and low acceptance, intention and willingness to get vaccinated. ^31,32^

COVID-19 vaccine hesitancy occurs against a backdrop of social-cultural complexities, poor government response in demystifying social and traditional myths and theories, and poor community involvement in public health measures. ^33^ Pregnant women in our study reported not having adequate information about the COVID-19 vaccine and were therefore concerned about the side effects of the vaccine on their pregnancy, vaccine safety, and efficacy, findings which were similar to an earlier study by Dinga et al. ^34^ In our study, religious affiliation was not associated with vaccine hesitancy, and neither was the need to seek permission for vaccination from a family member or partner.

Most unvaccinated pregnant women recruited into this study were willing to accept a COVID-19 vaccine if it were offered to them. However, they reported that a vaccine had not been offered to them, potentially pointing to insufficient effectiveness of mass media campaigns (which were widespread during this period in Zambia) and individual outreach during routine health care visits in demand creation. Pregnant women who expressed willingness to receive a vaccine on the day of their first ANC visit were referred to a designated center to receive a vaccine even though their intake was not documented by the study.

Several factors associated with low vaccine uptake were identified, including; age group, educational level, and HIV infection. In studies from other African countries, COVID-19 vaccination uptake has varied across age groups. ^32,35,36^ In our study, older pregnant women were more likely to have received a COVID-19 vaccine compared to younger women. This increase in uptake could be attributed to the government targeting older persons for receipt of the vaccine or increased understanding of risks and their prevailing knowledge of how the COVID-19 disease impacts older people given their own advancing age. ^26^ COVID-19 risk perception and behavior can be influenced by a higher level of education among populations. ^37^ While some studies have not observed any associations between educational level and COVID-19 vaccine uptake, ^38^ our findings contradict those found in Ethiopia where vaccine uptake was lower among participants who had a higher level of education and had attended college or university possibly due to their increased access to information and awareness of the potential side effects and safety. ^26,39^

Risk perception is a critical factor in vaccine decisions. ^40^ In this study, pregnant women who were not employed were less likely to receive a COVID-19 vaccine compared to those who were engaged in formal and informal work. This reduced uptake among the unemployed could be attributed to their perceived lowered risk of infection as they had limited contact with individuals outside of their household, particularly when lockdown regulations were strictly enforced. Additionally, a history of ever testing positive for COVID-19 influenced pregnant women’s decision to get a COVID-19 vaccine. These results corroborate with findings from studies in Indonesia and the UK which found that a higher perceived risk of COVID-19 infection was associated with higher acceptance of the vaccine. ^41–43^ Moreover, in Indonesia and the UK, receiving the vaccine was perceived to be vital for protecting oneself and preventing disease transmission.

Similarly, a higher proportion of pregnant women who had contact with a known or suspected COVID-19 patient within or outside their household were more likely to receive a COVID-19 vaccine. However, a majority of the pregnant women in this study indicated that they did not know about being in contact with a positive COVID-19 case. This could be because many cases of COVID-19 are asymptomatic or that symptoms are mild and nonspecific, and people did not get tested, or due to the limited availability of COVID-19 testing in Zambia. A household study in six districts in Zambia during the first wave estimated about one confirmed COVID-19 case for every 92 infections, ^44^, and in this study there continued to be substantial case ascertainment gaps. ^45^

COVID-19 vaccine hesitancy has been investigated among people living with HIV (PLWHIV). In Zambia, the MOH targeted PLHIV for COVID-19 vaccinations given their elevated risk of severe disease and substantial investments and achievements in HIV care in the country. ^46^ In our study, we assessed vaccine hesitancy among HIV-infected pregnant women and found a high degree of COVID-19 vaccine uptake among HIV-infected pregnant women as compared to those who were not infected with HIV. These results suggest high levels of vaccine acceptability and accessibility among pregnant women living with HIV. ^47^ Our results are similar to those reported in Nigeria where one in five women living with and at risk of HIV had a lower likelihood of being vaccine-hesitant. ^48^

For this study, data collected over 13 months shows that there was a modest increase in vaccination uptake among study participants from 0.4% at the beginning of the study in September 2021 to 2.6% at the end of the study in September 2022. This is in contrast to results from a meta-analysis that showed a higher rise in pooled COVID-19 vaccine acceptance rates in Ethiopia from 14.1% in 2022 to 42.46% in 2023. ^26,49^ Of note is that in this study, a proportion of the pregnant women who reported receiving a vaccine may have been vaccinated before they conceived. The gradual increase in the proportion of vaccinated pregnant women in our study could be attributed to an increase in the number of vaccine doses received by the MoH over time and their embarking on an ambitious 10-day COVID-19 vaccination campaign to reach the 70% eligible population in May 2022. ^50^

### Strengths and limitations of the study

A major strength of this study was the leveraging of infrastructure and human resources in the MNCH departments at public health facilities to enroll pregnant women seeking routine ANC services. ^51^ Health care providers and community-based volunteers, who were already providing ANC services at the study sites were engaged to assist with carrying out study procedures thereby easing the study implementation process, reducing costs, and gaining the confidence of participants. The study was able to access a longitudinal selection of pregnant women over time and triangulate a limited amount of routine data collected from part of the national DHIS2-COVAX tracker with data collected from study participants from participants to validate the findings. A limitation of the study is that the analysis does not include vaccine data of almost 1,837 participants as they were recruited into the study before the vaccine follow-up questions were appended to the questionnaire. Additionally, the districts were purposefully selected so the findings might not be generalizable to all of Zambia.

## CONCLUSION

Using pregnant women seeking routine antenatal care in public health facilities to assess COVID-19 vaccination status, uptake over time, intent, and acceptance of vaccine is acceptable and feasible in a low-resource setting. Overall, this study found a low uptake of COVID-19 vaccines among pregnant women attending first ANC in rural and urban settings, indicating a need to reinforce vaccine uptake efforts to prevent severe disease and adverse outcomes among these vulnerable populations. The leveraging of existing infrastructure and human resources provided an easy, sustainable platform for routinely monitoring COVID-19 vaccine, and consequently helped to understand vaccine hesitancy among pregnant women. This is critical for devising key vaccination messages and can facilitate the design and adoption of mass vaccination strategies for minority groups, reduce vaccine hesitancy, and subsequently increase vaccine uptake. Demand generation for vaccines might be more effective if done during routine health services and targeted at individuals (i.e., inviting pregnant women to get vaccinated). This would be possible by incorporating the vaccine as part of the routine ANC package to increase coverage among pregnant women.

## Data Availability

The authors confirm that the data supporting the findings of this study are available within the article. Raw data that support the findings of this study are available from the corresponding author upon request.

## Authors’ contributions

Conceptualization was done by JG, JH, IS, and TT. They also provided oversight for the implementation of study activities. Data curation was done by TT, BK, KK, and M-RT. Data analysis was guided by SB and performed by PS. The initial draft was prepared by TT. All authors proofread and approved the final version of the manuscript.

## Acknowledgments

The authors would like to acknowledge the Ministry of Health, Provincial Health Directors – Eastern and Lusaka provinces, District Health Directors – Chadiza, Chipata, Chongwe, and Lusaka Districts, and the National Health Research Authority for their collaboration at the preparatory stage of the study. We also wish to acknowledge the healthcare workers and community health workers (CHWs) at the health facilities for their participation and support during the implementation of study activities. Our special gratitude goes to the Research Assistants on the study – Beatrice Chibundi, Chikatizyo Mhango, Linda Phiri, and Patricia Z. Shabalu – for providing oversight for the implementation of study activities at each of the 39 health facilities.

## Disclaimer

The findings and conclusions in this paper are the products of study work implemented in Zambia and thus represent the opinions of the author(s). The paper does not necessarily represent the official position of the funding agency.

